# Subthalamic Nucleus Deep Brain Stimulation Alleviates the Sequence Effect and Freezing of Gait in Parkinson’s Disease

**DOI:** 10.1101/2025.05.08.25327015

**Authors:** Chuyi Cui, Goun Je, Kevin B. Wilkins, Tilman Schulte, Helen M. Bronte-Stewart

## Abstract

**Background:** The sequence effect in gait (progressive shortening of strides) contributes to freezing of gait (FOG), a debilitating feature of Parkinson’s Disease (PD). The sequence effect is refractory to dopaminergic medication or attentional strategies, with alleviation observed only from external cueing. While subthalamic nucleus (STN) deep brain stimulation (DBS) is a standard treatment for PD, its efficacy for gait impairment and FOG remains debated.

**Objective:** To characterize the sequence effect in spatial-temporal gait parameters in PD and investigate the effect of STN-DBS.

**Methods:** Eighteen individuals with PD with bilateral STN-DBS and fourteen age-matched healthy controls performed a harnessed stepping-in-place task, which we previously validated to elicit FOG. PD participants were assessed OFF and ON DBS in the off-medication state. Spatial-temporal gait parameters were obtained from inertial measurement units placed on both shanks, and the sequence effect was modeled with an exponential decay function for each gait parameter over time.

**Results:** The sequence effect was evident in shank swing angular velocity and range among PD participants, which was absent in healthy controls. The degree of sequence effect was significantly correlated with the time spent freezing during off therapy (*p* < 0.001). STN-DBS significantly alleviated the sequence effect (*p* = 0.002) and reduced the time spent freezing (*p* = 0.01).

**Conclusion:** The stepping-in-place task elicited the sequence effect in both velocity and spatial aspects of gait in individuals with PD. The sequence effect contributed to FOG severity. STN-DBS was effective in alleviating the sequence effect, thereby improving FOG in PD.

## Introduction

The sequence effect, a phenomenon recognized specifically in Parkinson’s Disease (PD)^1,2^, is characterized by a progressive deterioration in the quality of successive motor actions, which manifests in various movements, including upper-limb tasks, speech and gait^3–5^. In gait, the sequence effect is often evident as a gradual step-to-step reduction of step length. This gait deficit has been identified as a major contributor to freezing of gait (FOG), a deliberating symptom of PD that increases the risk of falls^3,6–10^. This underscores the importance of understanding the sequence effect for the effective management of gait disturbances in PD.

The sequence effect results from underlying deficits in the maintenance of specified motor parameters and cue production within basal ganglia^8,11^. It is largely unresponsive to dopaminergic medication or attentional strategies^2,3,12^, with alleviation observed only from external visual or audio cueing^3,13,14^. Deep brain stimulation (DBS), specifically targeting the subthalamic nucleus (STN) or globus pallidus interna (GPi), is a well-established treatment for motor symptoms in PD including tremor, rigidity, and bradykinesia. However, its efficacy in addressing gait impairment, the sequence effect during gait, and FOG has been subject to debate^15,16^. Further research is needed to quantitatively evaluate gait characteristics to better understand the effect of DBS therapy on gait dysfunction in PD.

We recently demonstrated the potential of the STN-DBS to alleviate the sequence effect in upper-limb bradykinesia in individuals with PD^17^. Building on these findings, the present study aimed to investigate the impact of STN-DBS on the sequence effect during gait and furthermore on FOG. We employed a repetitive stepping-in-place (SIP) task, which we previously validated to elicit and measure FOG. The SIP task controls for potential confounding influences of optic flow or external cues on gait by enclosing the participant in a stationary visual surround. The goal of this study was to examine the presence of the sequence effect in spatial-temporal gait parameters in individuals with PD, in comparison to a group of age-matched healthy controls, and how the sequence effect may be alleviated by STN-DBS.

## Methods

### Participants

Participants included 18 individuals with clinically established PD (six females, age: 61.2 ± 8.4 yrs) and 14 healthy controls (seven females, age: 62.8 ± 7.1 yrs).

PD participants were chronically implanted with bilateral DBS omnidirectional leads (Legacy, 3389, Medtronic, Inc.) in the sensorimotor region of STN, which were connected to an investigative implantable pulse generator (IPG), either Activa™ PC+S, or Summit RC+S (Medtronic, Inc.). All patients met pre-operative selection criteria (see Supplementary Information). The surgical techniques have been previously described^18^. Additional inclusion criteria for the present study included being able to ambulate in the off-medication OFF-DBS state. PD patients provided written informed consent, which was approved by the Food and Drug Administration with an Investigational Device Exemption and by the Stanford University Institutional Review Board.

Healthy control participants were at least 45 years of age. Exclusion criteria included fewer than eight years of education, a history of psychiatric (e.g. schizophrenia or bipolar disorder), neurological, or medical condition (e.g. stroke, diabetes) potentially affecting the central nervous system, or cognitive score less than Dementia Rating Scale (DRS-2) cut-off score (136/144)^18^.

All healthy control participants provided written informed consent, which was approved by the Institutional Review Board of Stanford University and the SRI International.

### Gait Task

During the SIP task, the participant performed alternating stepping on a dual dynamic force plate system, with one foot on each plate, in an enclosed environment that minimized external sensory information (Neurocom Inc., Clackamas, OR, USA; Bertec Corporation, Columbus, OH, USA). The task consisted of 30 seconds of quiet stance followed by 100 seconds of stepping. Upon a “Go” cue, the participants began raising their legs alternately at a self-selected pace while harnessed. Participants were instrumented with Inertial Measurement Units (IMUs, APDM, Inc., Portland, OR) attached to bilateral shank segments (lower legs).

### Experiment Protocol

All PD participants were clinically optimized on DBS: the experiment was conducted between six months and five years after their initial DBS programming. PD participants were off dopaminergic medication, which entailed the withdrawal of long-acting dopamine agonists for 48 hours, dopamine agonists and controlled release carbidopa/levodopa for 24 hours, and short acting medication for 12 hours prior to the study visit. Each PD participant performed one trial of SIP OFF DBS and one trial ON DBS. For the OFF DBS condition, stimulation was turned off for at least 15 minutes before participants performed the gait task to wash out the stimulation effect^19^. For the ON DBS condition, participants performed the task with DBS settings at the clinical or clinically equivalent stimulation intensity, with a stimulation frequency of 140 Hz or 150 Hz, and a pulse width of 60 µs.

### Data Acquisition

Kinetic data was obtained from the dual force plates, sampled at either 100 or 1000 Hz. Kinematic data was measured from the IMUs, sampled at 128 Hz, and synchronized with each other in MotionStudio (APDM, Inc., Portland, OR, USA). Kinetic and kinematic data were synchronized by a rapid hammer strike event on one of the force plates, which was captured concurrently by both kinetic and kinematic signals.

### Data Analysis

The vertical ground reaction forces (GRF) under each foot were low-pass filtered using a zero-lag second order Butterworth filter with 12 Hz cutoff frequency and normalized to body weight. FOG episodes were identified from the vertical GRF time series using a previously validated algorithm^20^. The percent time spent freezing was calculated as the total duration of freezing over the 100-second task, expressed as a percentage (0-100%).

The tri-axial gyroscope signals from each shank IMU sensor were low-pass filtered using zero-lag 8th order Butterworth filters with a 9 Hz cutoff frequency. The tri-axial signals were preprocessed through principal component analysis to extract the 1-D angular velocity aligned with plane of the shank swing. Positive angular velocity represents the leg lift motion and shank swinging backward during the first half of the single stance phase, while negative angular velocity corresponds to the shank moving down towards ground during the second half of the single stance (Figure 1A,B).

**Figure 1.**
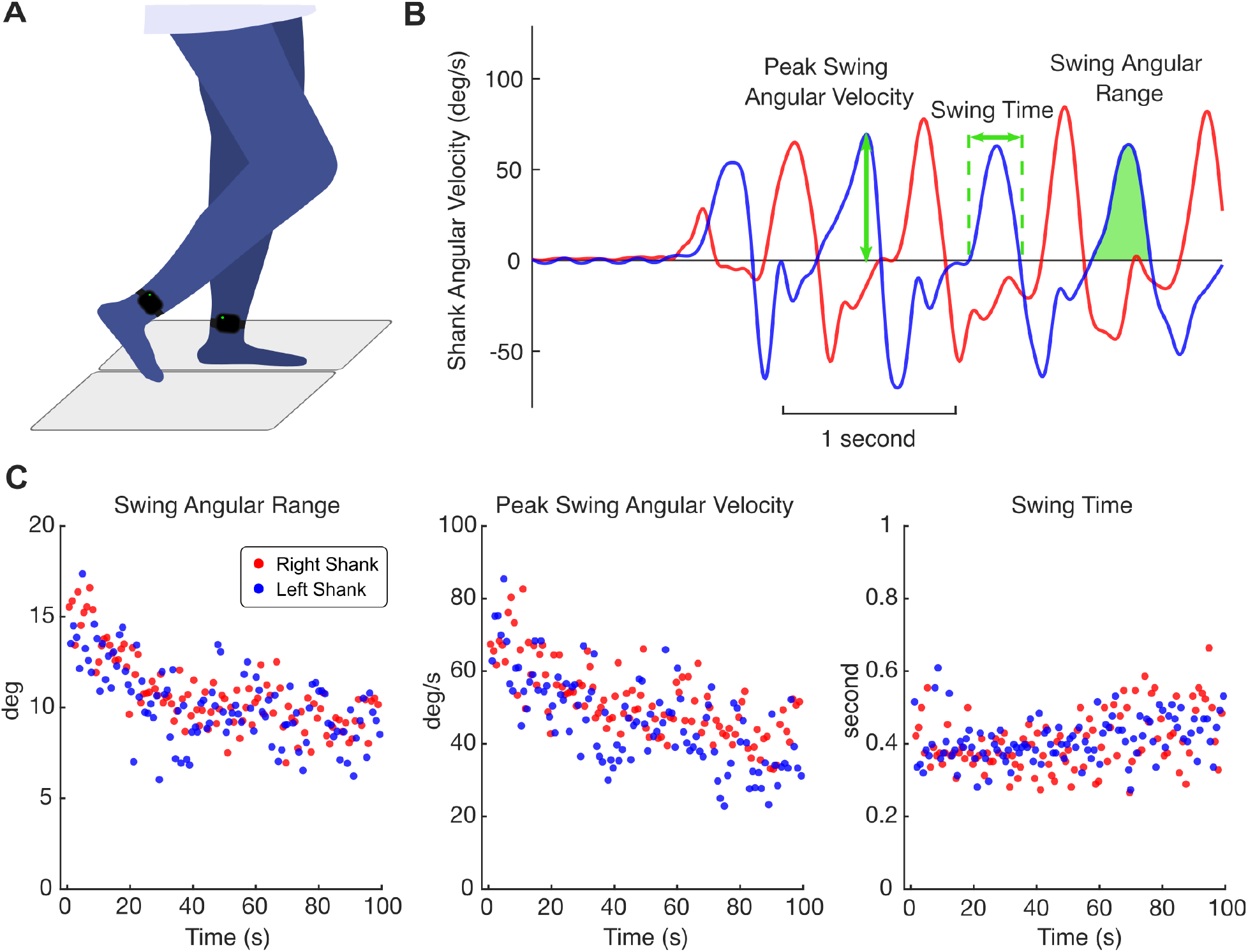
(A) Side view depiction of a participant performing the stepping-in-place (SIP) task, with one foot on each force plate (gray), wearing inertial measurement units (IMUs) on both shanks. (B) Time series of shank angular velocity (AV) in the sagittal plane. Peak swing AV, swing time, and swing angular range parameters were identified from the shank AV time series. (C) Extracted gait parameters over the 100 second stepping for an example PD participant off medication and OFF DBS.

Three gait parameters were obtained from the angular velocity time series of each shank: peak swing angular velocity, swing time, and swing angular range. Shank angular velocity peaks were detected with a threshold of minimum peak height of 10 deg/s. Each peak represents the maximum swing velocity during the leg lift phase of a step. Swing time is calculated as the interval between the two zero-crossings before and after each identified peak, which was the time taken from the initial shank swing to the maximum swing height. Lastly, the shank angular range was calculated as the integral area under the angular velocity curve during the identified swing time (Fig. 1B).

To quantify the sequence effect, we fitted a curve to each of the three extracted gait parameters over time. Given the asymmetrical motor symptoms in PD, we assessed the sequence effect for each shank separately. Gait deterioration was modeled using an exponential decay function:

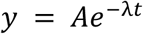

where *t* represents time, *y* is the respective gait parameter, and the decay rate *λ* describes the sequence effect. A larger *λ* value indicates a steeper decay over time, thereby more severe sequence effect (i.e. worse gait).

As some trials displayed multiple movement epochs of the sequence effect where the participant was able to reset following initial period of gait deterioration, all trials were examined to determine if the sequence effect should be quantified from the entire trial or within epochs. For trials with FOG episodes identified, the movement epoch before the start of the first freezing episode was isolated and used to quantify the sequence effect. For trials without FOG and with more than one epoch of the sequence effect, the first epoch was used, as had been done for limb bradykinesia^20^.

### Statistics

Statistical analyses were conducted in Python. Linear mixed effects models were used to test the difference between the healthy control and PD groups for each sequence effect metric with a fixed effect of group and random effect of participant nested with body side. Linear mixed effects models were performed within the PD group to examine the effect of DBS on each sequence effect metric using a fixed effect of stimulation condition and random effect of participant nested with body side. The effect of DBS on the percent time freezing were evaluated using Wilcoxon signed-rank test given the non-normal distribution of data. Lastly, to explore the relationship between the sequence effect and FOG symptom, Spearman’s rho correlations were performed between each sequence effect metric and the percent time freezing across PD participants OFF DBS. Significance was set at α < 0.05 for all tests.

## Results

Demographic information for the PD cohort is shown in Table 1. PD participants had an off medication/OFF DBS MDS-UPDRS III score of 36.1 ± 10.9. The PD cohort and the healthy control cohort were age matched (t = 0.84, *p* = 0.40).

**Table 1.** PD cohort demographics.

| Patient No. | IPG | Sex | Age (years) | Disease Duration (years) | MDS-UPDRS III Total Score | Clinical Phenotype |
| --- | --- | --- | --- | --- | --- | --- |
| 1 | Activa | M | 56-60 | 8 | 47 | Non-freezer |
| 2 | Activa | M | 61-65 | 7 | 46 | Freezer |
| 3 | Activa | M | 56-60 | 8 | 42 | Freezer |
| 4 | Activa | M | 41-45 | 8 | 34 | Freezer |
| 5 | Activa | M | 56-60 | 10 | 54 | Freezer |
| 6 | Activa | M | 71-75 | 11 | 43 | Non-freezer |
| 7 | Activa | F | 61-65 | 13 | 36 | Freezer |
| 8 | Activa | F | 66-70 | 9 | 9 | Non-freezer |
| 9 | Activa | F | 51-55 | 13 | 41 | Freezer |
| 10 | Activa | M | 61-65 | 20 | 29 | Freezer |
| 11 | Activa | F | 56-60 | 15 | 36 | Freezer |
| 12 | Activa | M | 56-60 | 13 | 29 | Freezer |
| 13 | Activa | M | 46-50 | 12 | 36 | Freezer |
| 14 | Summit | M | 61-65 | 11 | 43 | Non-freezer |
| 15 | Summit | F | 71-75 | 17 | 19 | Freezer |
| 16 | Summit | M | 66-70 | 13 | 34 | Freezer |
| 17 | Summit | M | 71-75 | 16 | 26 | Freezer |
| 18 | Summit | F | 56-60 | 15 | 45 | Freezer |

Note: The Movement Disorders Society Unified Parkinson's Disease Rating Scale Part III (MDS-UPDRS III) Score was obtained in the off-medication OFF-DBS state. Clinical phenotypes were classified based on the clinical history of a patient's FOG symptom.

### Sequence Effect and the Effect of DBS

Among PD, a total of 20 out of 36 trials demonstrated multiple movement epochs, including 18 trials where participants had FOG, and two trials where participants showed a reset following the initial decay of swing angular velocity (Supplementary Figure 1). In the other sixteen trials, the participants were able to perform SIP in continuous manner, therefore the sequence effect was analyzed over the full 100-second duration.

Figure 2 shows representative ground reaction forces, kinematics, and sequence effect for a healthy control participant and individual with PD both OFF DBS and ON DBS. The healthy control maintains consistent stepping throughout the 100 seconds with minimal sequence effect. Meanwhile, OFF DBS, the individual with PD shows a notable sequence effect characterized by a decrease in swing angular velocity that culminates in a freeze approximately 30 seconds into the task. However, ON DBS, the participant exhibits no freezing and is able to maintain consistent stepping throughout with minimal sequence effect.

**Figure 2.**
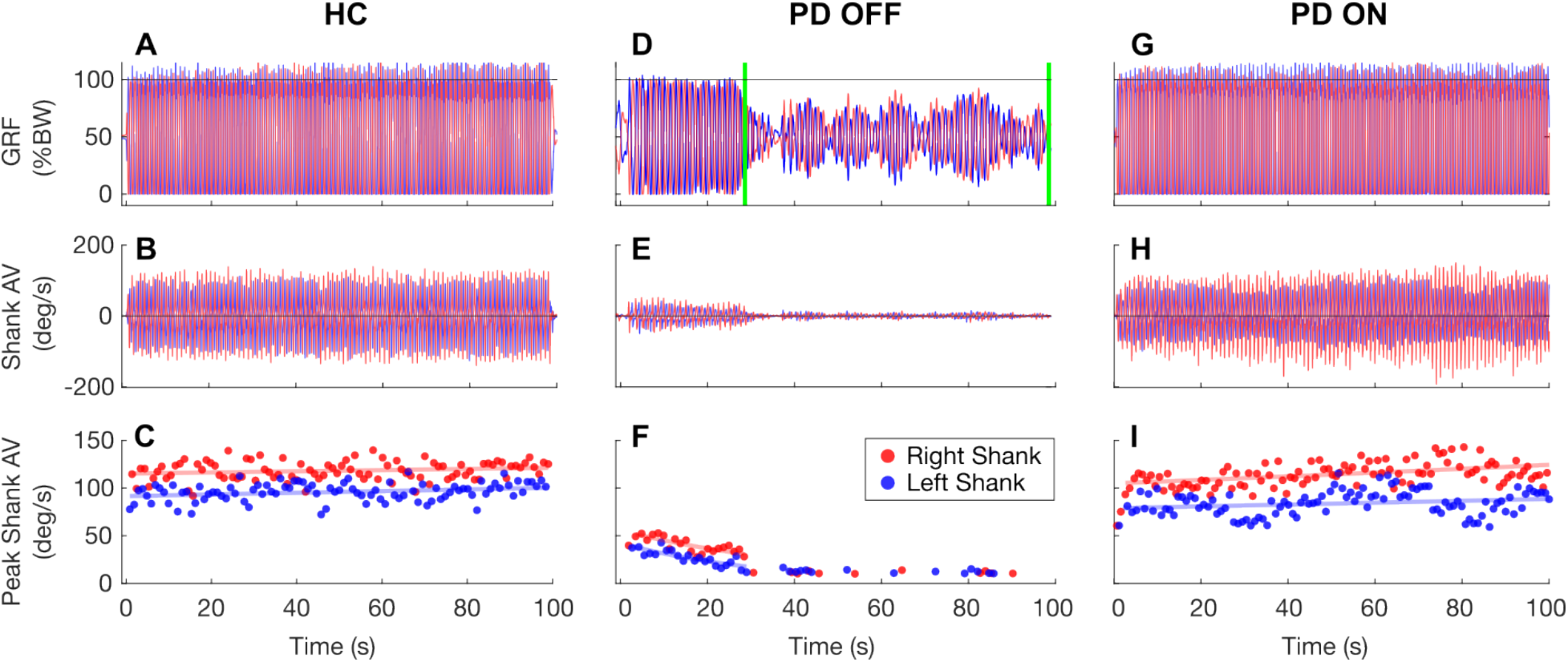
Example SIP task performance from one healthy control participant (A-C) and one PD participant (D-I) OFF and ON DBS. (A,D,G) Time series of vertical ground reaction forces (GRF) under left (blue) and right (red) foot, expressed as the percentage of the participant’s body weight. (B,E,H) Time series of the shank angular velocity measured from the left (blue) and right (red) shank IMU sensors. (C,F,I) The peak swing angular velocity for each identified step. An exponential curve was fit over this metric for each leg, respectively. (D) Green lines mark a freezing episode identified from the GRFs by the computerized algorithm. (E,F) The first movement epoch before the start of the freezing episode was isolated for quantifying sequence effect.

Figure 3 depicts the sequence effect (*λ*) quantified for each of the three gait parameters for healthy control and PD groups. Linear mixed models showed that the PD group, OFF DBS, had significantly greater sequence effect in peak angular velocity (β = 0.010, 95% CI [0.006, 0.014], *p* < 0.001) and swing angular range (β = 0.008, 95% CI [0.003, 0.013], *p* = 0.001) compared to age-matched healthy control. There was no significant difference for the sequence effect in swing time between the two cohorts (β = 0.001, 95% CI [−0.002, 0.004], *p* = 0.727).

**Figure 3.**
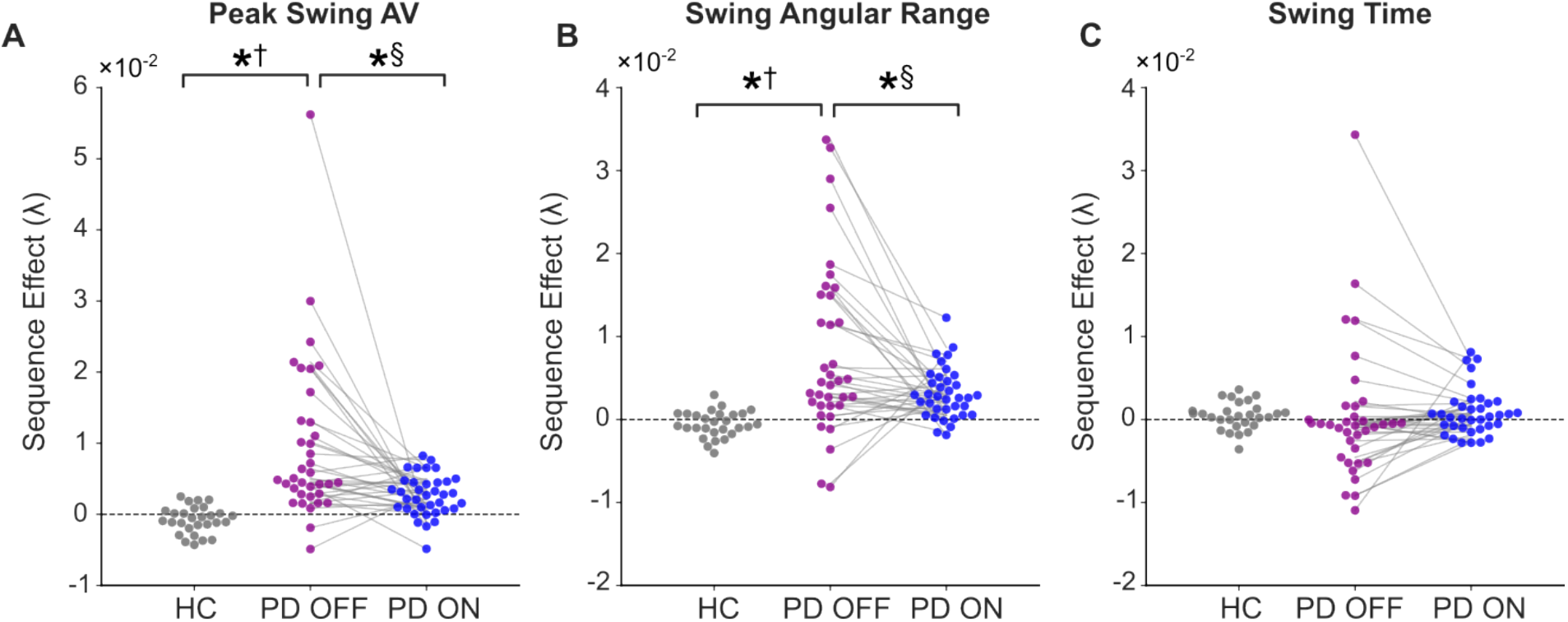
Sequence effect metrics comparing healthy controls (N=14) and PD participants (N=18) in OFF and ON DBS states. Sequence effect (λ) was quantified for each of the three gait parameters: peak swing angular velocity (A), swing angular range (B), and swing time (C). Each dot represents one leg of a participant, with grey lines connecting the OFF DBS and ON DBS states for each PD participant’s body side. The horizontal line indicates no change over time. Positive values reflect decay, while negative values indicate growth. Asterisks denote significant differences: † indicates significant group differences between healthy controls and PD participants OFF therapy, and § indicates significant effects of DBS within PD group.

Fig. 3 also depicts the sequence effect (*λ*) ON versus OFF DBS within the PD group. Linear mixed models showed that DBS significantly reduced (improved) the sequence effect in peak angular velocity (β = −0.007, 95% CI [−0.010 −0.003], *p* < 0.001) and swing angular range (β = −0.005, 95% CI [−0.008, −0.002], p = 0.002) compared to the OFF DBS condition. There was no significant difference in the sequence effect in swing time between stimulation conditions (β = 0.001, 95% CI [−0.002, 0.003], *p* = 0.620).

### Sequence effect and FOG interactions

Twelve PD participants exhibited FOG episodes during the SIP task, as identified from the GRF time series. The Wilcoxon signed-rank test showed that the percent time freezing was significantly reduced ON DBS compared to OFF DBS (W = 6, *p* = 0.010, Figure 4A).

**Figure 4.**
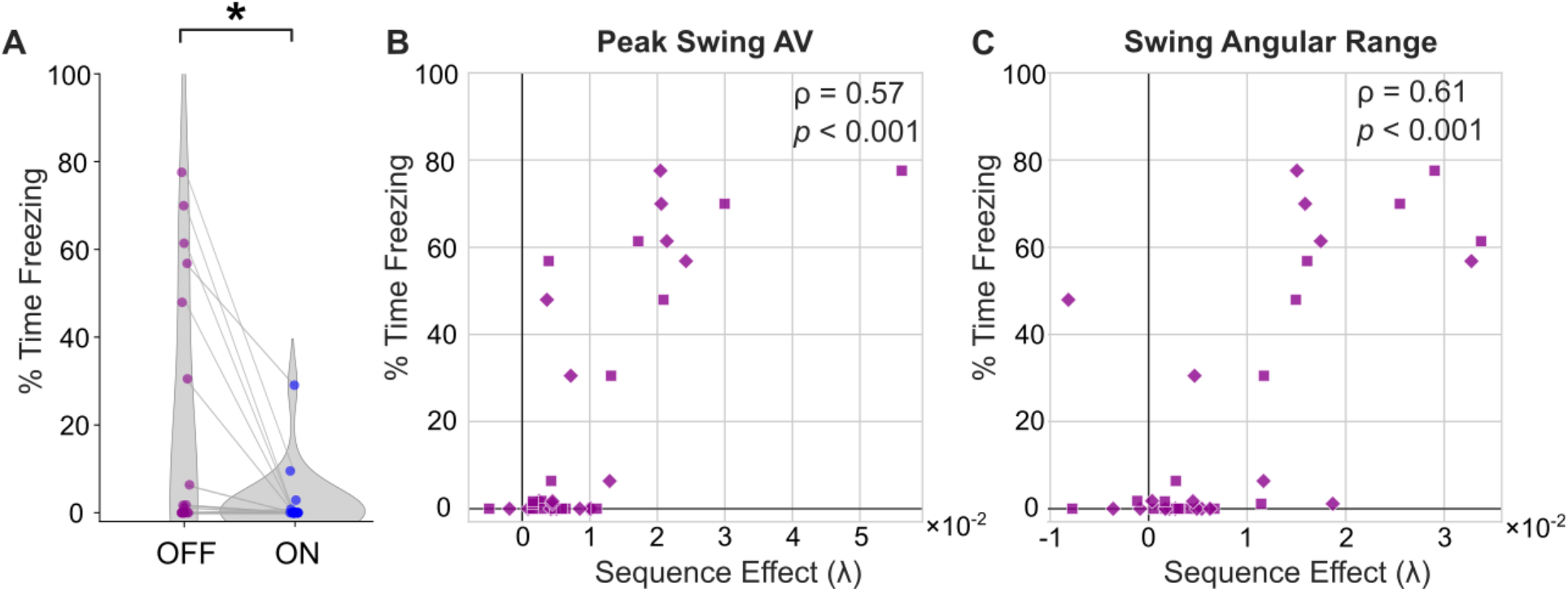
(A) The percent time freezing during SIP OFF versus ON DBS for PD participants. Asterisk denotes significant effect of DBS. (B, C) Correlation between the sequence effect and the percentage of time spent freezing OFF DBS. Each point represents one body side of each participant, with different marker symbols used to distinguish between the left and right legs.

Spearman’s rho analysis for the OFF DBS condition showed significantly strong correlations between the sequence effect in swing angular velocity and the percent time freezing (ρ = 0.57, *p* < 0.001, Fig. 4B), and between the sequence effect in swing angular range and percent time freezing (ρ = 0.61, *p* < 0.001, Fig. 4C), with greater sequence effect correlated with longer total time spent in freezing episodes.

## Discussion

The present study investigated how the sequence effect manifested in a SIP task in a cohort with moderate to severe PD in comparison to a cohort of healthy controls and evaluated the efficacy of STN-DBS on the sequence effect in PD. We demonstrated that the sequence effect in PD off therapy was evident in velocity and spatial domains of SIP and confirmed its role in contributing to FOG. STN-DBS significantly alleviated the sequence effect and reduced freezing severity.

### SIP task elicits the sequence effect in PD

The sequence effect in gait has been mainly investigated during overground walking, and described as the progressive reduction of stride or step length^3,6,9,21,22^. However, FOG is generally difficult to evaluate during open space walking such as in the clinic or in laboratory settings ^23–25^. Moreover, environmental cues or constraints may modify gait: congruent optic flow assists gait while short walkways limit observation to ‘destination sequence effect’^21,22^. Our present study leveraged the repetitive SIP task, validated to elicit FOG^20^, and examined the gait kinematics in the period prior to the occurrences of freezing events. The use of an enclosed visual surround offered the opportunity to observe potential gait deterioration over a relatively long duration (i.e., 100 s) of continuous SIP without the effect from external visual cues.

One of the main findings was the significant reduction of both leg swing angular velocity and angular range over time in the PD cohort when off therapy (Fig. 3 A, B), revealing how sequence effect manifested in gait in PD. This gait sequence effect was absent in aged-matched healthy controls, underscoring the PD-specific motor deficit. This deficit is believed to stem from dysfunction within the basal ganglia circuitry in sensory motor integration and cue production, which lead to a mismatch between the intended and performed movement^8,11^.

Interestingly, SIP swing time demonstrated variable trends across PD participants: some patients displayed a decrease of swing time (*λ* > 0) over time, while some others showed an increase (*λ* < 0, Fig. 3C). This revealed two distinct types of gait deterioration observed among individuals. In one scenario, the sequence effect occurred primarily in step amplitude (i.e. smaller swing angular range) than in speed, leading to a decrease of swing time over time. In the other scenario, the sequence effect occurred primarily in speed (i.e. slower swing velocity), resulting in an increase of swing time observed. Additionally, we observed that stable swing time (i.e., *λ* close to zero) does not necessarily indicate consistent stepping performance. Instead, it may reflect a proportional decay in both velocity and amplitude (Fig. 1). Since either variable or consistent swing time could be associated with actual gait deterioration, our findings suggest that the temporal aspect alone is not a good indicator of the sequence effect in gait, which is more reliably reflected in velocity and spatial aspects.

### Sequence effect predicts FOG severity

There was a significant correlation between the degree of the sequence effect (*λ*) from the first epoch and the severity of FOG (percent time freezing) obtained from the entire trial (Fig. 4 B,C). This suggests that the more rapid the deterioration of speed and/or amplitude the greater the difficulty to maintain forward progression, and the more likely it will lead to freezing events. This aligns with the existing knowledge about how the sequence effect contributes to FOG^3,6^. More specifically, we observed a cluster of individuals who exhibited more gradual deterioration in SIP velocity and amplitude (milder sequence effect), which was not associated with freezing episodes. Given that FOG is a progressive symptom typically emerging later in the course of the disease ^26^, the sequence effect observed in gait kinematics could potentially act as an early marker for the development of FOG symptoms. Our PD cohort primarily consists of freezers, with only four clinical non-freezers (Table 1). Due to the small sample of non-freezers, we were unable to statistically confirm whether the sequence effect is present even in PD non-freezers. However, descriptively, two out of the four PD non-freezers, while off therapy, exhibited greater decay in swing angular velocity than the maximum observed among healthy controls; a similar trend was observed in swing angular range (Supplementary Fig. 2). Future research involving a larger sample and longitudinal follow-up is needed to further confirm this.

### STN-DBS alleviated sequence effect and reduced FOG

There is a mixed body of literature regarding the efficacy of DBS on gait impairment and FOG^15,16^. A widely held view posits that DBS primarily addresses symptoms such as tremor, bradykinesia, and rigidity more effectively than axial symptoms in PD. In the present study, we observed that STN-DBS significantly reduced the sequence effect, demonstrated as slower decay of the swing velocity and amplitude over time. Meanwhile, the percent time freezing also reduced ON STN-DBS. Our findings add to the evidence supporting that STN-DBS improves gait and FOG symptoms by mitigating the sequence effect manifest in step velocity and amplitude in PD.

The discrepancy in the literature regarding the efficacy of DBS on gait impairments and FOG could be attributed to a multitude of factors associated with FOG and FOG subtypes. One important aspect of FOG is that it may be triggered by specific environmental contexts such as approaching a narrow doorway and turning^23^. The protocol in the present study does not fully account for scenarios such as gait-initiation or turning freezes. Moreover, nonmotor factors such as anxiety^27,28^ and attention^29^ also play roles in FOG. Recent imaging studies point to additional nondopaminergic mechanisms that contributes to gait variability and FOG such as changes in cholinergic system^30,31^ and noradrenergic limbic pathways^32^ in individuals with PD, which are typically not well-targeted by STN-DBS. Collectively, these factors may limit the overall efficacy of STN-DBS in treating FOG and influence which patient phenotypes are most likely to benefit from this intervention.

## Limitations

While the SIP task offers advantages to investigate the sequence effect, it has potential limitations. Notably, it is a less rehearsed motor activity, and there is no explicit ‘task goal’ to maintain consistent movement speed and amplitude as naturally controlled for during overground walking. These factors may contribute to increased gait variability. To address this, we included a cohort of age-matched healthy controls. No significant growth or decay was observed in healthy controls, suggesting that the decay observed in PD in the present study reflects basal ganglia deficit rather than an artifact of task difficulty or peripheral fatigue.

Another limitation of the present study is that we only quantified the first epoch, despite that some trials could display multiple epochs of sequence effect (Supplementary Fig. 1). This approach was adopted to mitigate variability in the number of movement epochs across participants and conditions, thus avoiding confounding the statistical analysis. Additionally, only the first epoch allows for a comparable motor plan across all groups. Future research could examine all sequences to understand the neuromotor control of the movement reset following the initial decay and how that potentially contributes to gait function in individuals with PD.

## Conclusion

In this study we demonstrated the repetitive stepping-in-place task elicits the sequence effect in both velocity and spatial aspects of gait in individuals with PD, and that the degree of sequence effect correlated with the severity of the FOG. STN-DBS is effective in alleviating the gait sequence effect thus improving FOG symptom in PD.

## Data Availability

All data produced in the present study are available upon reasonable request to the authors

## Acknowledgment

The authors would like to thank the participants who dedicated their time to this study. This work was supported by NINDS UH3 NS107709, UH3 NS128150, R21 NS096398, NIAAA R01 AA023165, NIA R01 AG081144, Michael J. Fox Foundation (9605), Robert and Ruth Halperin Foundation, John A. Blume Foundation, John E. Cahill Family Foundation, and Medtronic Inc. who provided the devices used in this study but no additional financial support.

