## Supplemental Materials for "Subthalamic Nucleus Deep Brain Stimulation Alleviates the Sequence Effect and Freezing of Gait in Parkinson’s Disease"

**Inclusion and exclusion criteria**

For Activa™ PC+S cohort (NCT01990313), preoperative inclusion criteria included being at least 18 years of age; documented improvement in motor signs on versus off dopaminergic medication, the presence of complications of medication such as wearing off signs, fluctuating responses and/or dyskinesias, and/or medication refractory tremor, and/or impairment in the quality of life on or off medication, and being able to return for follow-up visits. Exclusion criteria for participants was dementia, untreated psychiatric disease, Hoehn and Yahr stage 5 (non-ambulatory) on medication, age greater than 80 years old, major surgical morbidities, such as severe hypertension, coagulopathy, and certain metabolic conditions that might increase the risk of hemorrhage or other surgical complications, presence of a cardiac pacemaker/defibrillator, and inability to understand/sign consent forms. For the Summit RC+S cohort (NCT04043403), preoperative selection criteria included being at least 18 years of age, meeting criteria for STN DBS, presence of complications of medication such as wearing off signs, fluctuating responses, dyskinesias, medication refractory tremor, and/or impairment in the quality of life on or off medication, and a score 1 on the Freezing of gait questionnaire and/or gait sub-score (Item 3.10) MDS-UPDRS III. Exclusion criteria included being over the age of 80, dementia, untreated psychiatric disease, Hoehn and Yahr stage 5, major surgical morbidities such as severe hypertension, coagulopathy, or conditions that might increase the risk of hemorrhage or other surgical complications, presence of a cardiac pacemaker, required rimes, ECT, MRI, or diathermy, pregnancy, cranial metallic implant, or history of seizures or epilepsy.

**DBS settings**

Clinical equivalent intensity was used for two Activa™ PC+S patients and all six Summit RC+S patients: For Activa™ PC+S patients, clinically equivalent DBS intensity was determined by observing similar therapeutic benefit to motor performance to what observed with the clinical DBS electrode configuration and intensity but in a sensing friendly configuration (Kehnemouyi et al., 2023). For Summit RC+S patients, the clinical equivalent stimulation intensity was selected to match the total electrical energy delivered of the patient’s calibrated adaptive DBS, which often resulted in slightly lower than their clinical intensity (Wilkins et al., 2024).

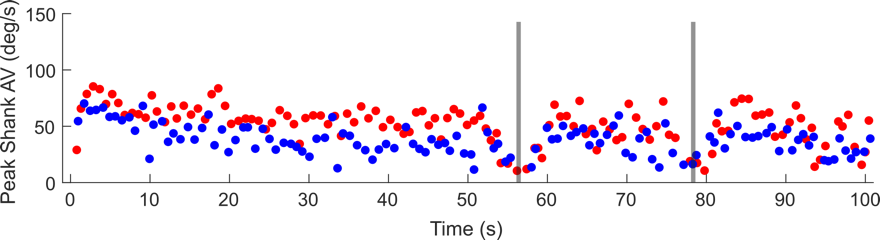

**Supplementary Figure 1**. Example SIP task with multiple movement epochs.

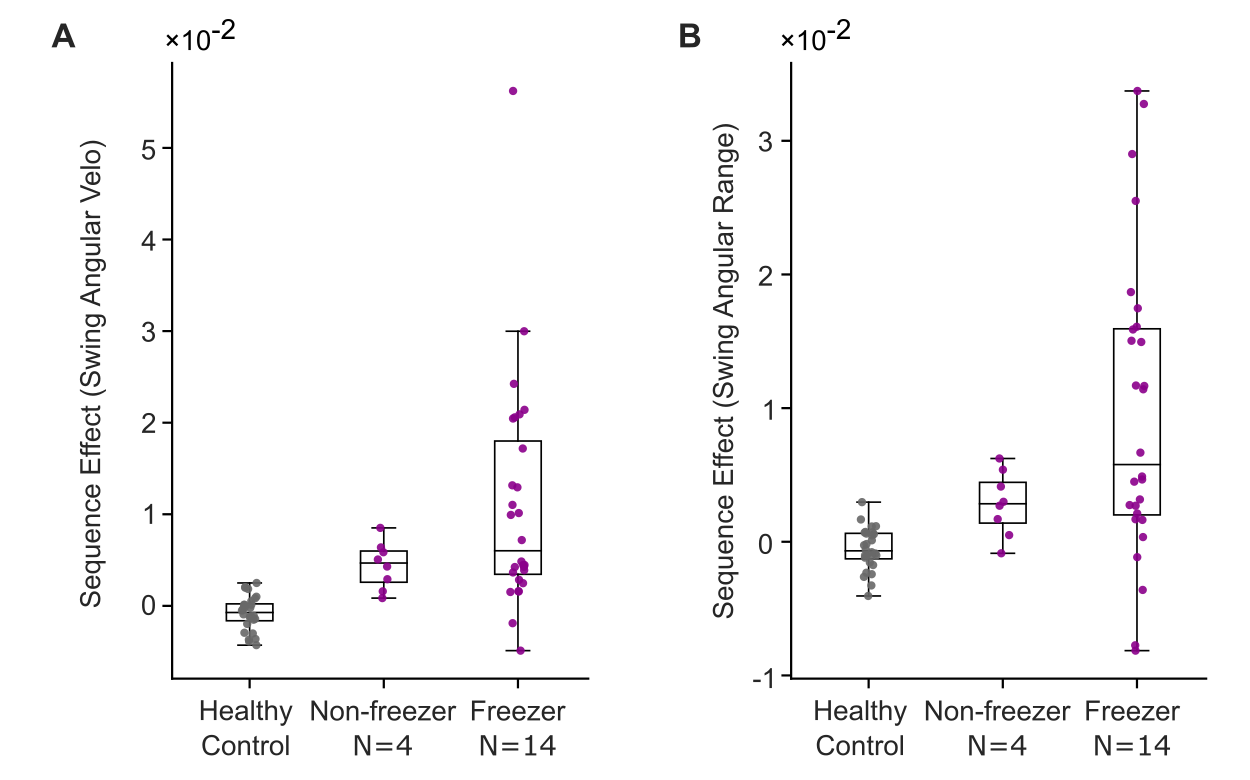

**Supplementary Figure 2**. Sequence effect of swing angular velocity (A) and swing angular range (B) across healthy control, PD nonfreezers, and PD freezers.

Kehnemouyi, Y. M., Petrucci, M. N., Wilkins, K. B., Melbourne, J. A., & Bronte-Stewart, H. M. (2023). The Sequence Effect Worsens Over Time in Parkinson’s Disease and Responds to Open and Closed-Loop Subthalamic Nucleus Deep Brain Stimulation. *Journal of Parkinson’s Disease*, *13*(4), 537–548. https://doi.org/10.3233/JPD-223368

Wilkins, K. B., Petrucci, M. N., Lambert, E. F., Melbourne, J. A., Gala, A. S., Akella, P., Parisi, L., Cui, C., Kehnemouyi, Y. M., Hoffman, S. L., Aditham, S., Diep, C., Dorris, H. J., Parker, J. E., Herron, J. A., & Bronte-Stewart, H. M. (2024). Beta Burst-Driven Adaptive Deep Brain Stimulation Improves Gait Impairment and Freezing of Gait in Parkinson’s Disease. *medRxiv*, 2024.06.26.24309418. https://doi.org/10.1101/2024.06.26.24309418
